# Temperature-related disasters in Europe – a cross-sectional analysis of the emergency events database from a pediatric perspective

**DOI:** 10.1101/19012633

**Authors:** Heiko Brennenstuhl, Manuel Will, Elias Ries, Konstantin Mechler, Sven Garbade, Markus Ries

**Author notes:** address correspondence to Markus Ries, MD PhD MHSc FCP, Pediatric Neurology and Metabolic Medicine, Center for Pediatric and Adolescent Medicine, University Hospital Heidelberg, Im Neuenheimer Feld 430, D-69120 Heidelberg, Germany.

## Abstract

This study investigates patterns of extreme temperature-related events in Europe and its significance for the public health, with a focus on the vulnerable pediatric population. A generalized additive model of average surface temperature development for the European countries is described and discussed with an in-depth analysis of the influence of temperature on evolutional and behavioral aspects.

**Methods:** Extreme temperature related events are recorded in the publicly available epidemiological database of Emergency Events (EM-DAT). A comparative and descriptive statistical analysis of this data was conducted with a focus on (prospective) records from 1988 onwards. Average surface temperature data was provided by the World Bank’s Climate Change Knowledge Portal. The criteria for strengthening the reporting of observational studies in epidemiology (STROBE) were respected.

**Results:** Within EM-DAT, extreme temperature-related disasters in Europe were categorized as either heat waves, drought, forest or land fires, or cold waves and severe winter conditions, accordingly. The most frequent type of event recorded were cold waves (36.2%). However, cold waves and severe winter conditions only accounted for about 6,460 casualties (4.4%), while heat waves were responsible for a total of 137,533 casualties (95.1%). During the prospective observational period of the EM-DAT database, heat waves in 2003, 2006, 2010, and 2015, claimed a total of 119,760 casualties. These most severe heatwaves were geographically distributed over Russia (2010), as well as France, Italy, Spain, and Germany, each in 2003. Accordingly, analysis of temperature data revealed an increasing average surface temperature for all assessed European countries, correlating with in an increasing frequency of extreme temperature-related events.

**Conclusion:** This study shows that according to EM-DAT data extreme temperatures are an increasingly important public health threat to the European population as the average European surface temperatures are rising. Although cold waves are more frequently reported in EM-DAT, heat waves are the major cause for temperature-related casualties. Therefore, we conclude that evolutional and cultural resilience against cold and drought is significantly higher than it is against heat. Our results project that the frequency, duration and intensity of heat waves will further increase due to current climatic changes and become a more prevalent problem for future generations. Hence, we propose an emergency plan to inform the public and authorities about measurements to be taken in such extreme heat conditions to overcome the prevailing lack of information available to the public.

## Introduction

According to NASA’s climate research, the average global surface temperature has increased by 1 °C relative to 1951-1980 average temperatures. Since 2001, 18 of the 19 warmest summer periods occurred with 2016 being the warmest on record. (Data source: NASA’s Goddard Institute for Space Studies (GISS)).^1^ As climate experts expect this trend to progressively continue, periods of extreme heat and cold will become more frequent in the future.

Exposure of humans to extreme temperatures causes a considerable danger to their health, with younger children and elderly people being amongst the most vulnerable populations. The average human body temperature is defined as 36 to 37.5 °C (96.8 to 99.5 °F). A deviation of ± 3 °C (5.4 °F) can overexert the body’s compensation mechanisms and cause substantial damage leading, ultimately, to death. Mechanisms to regulate body temperature include conduction and convection, radiation, and evaporation, the latter mostly achieved through peripheral cutaneous vasodilatation. While evaporation is the most effective way to reduce body temperature, it becomes ineffective as soon as relative humidity rises above 75%.^2^

Several characteristics contribute to the susceptibility of the pediatric population: Young children lack the ability to properly regulate body temperature and are at the same time in constant need of parental care and supervision of fluid replenishment. A higher surface area to mass ratio of children is responsible for increased absorption of heat.^3^ Additionally, children display a higher metabolic turnover rate, which is further enhanced by high temperatures, resulting in increased oxygen consumption with hyperpnea and tachycardia.^4^ Within the European Union, 15.6 % of the population, accordingly 115 million people, are currently below the age of 14 years (Data source: Eurostat; accessed July 11, 2019), representing a substantial and potentially vulnerable population in case of catastrophic temperature-related events.

According to a recent analysis of disaster patterns in Germany and France – the only of its kind up to date –, environmental temperature is an important mechanism of injury leading to disasters. Heat waves occurred less frequently in both countries compared with cold waves and severe winter conditions, but heat waves caused significantly more casualties in both countries.^5^ On this basis we hypothesize that population resilience in heat-related disasters is lower than in cold-related disasters leading to significantly higher casualties. In order to test this hypothesis, we directed. Here we our efforts towards investigating the epidemiological vulnerability of a larger, pan-European population related to specific climatological and meteorological conditions as reported in the Emergency Events Database.^6^ We statistically analyzed cumulated temperature associated events, casualties, and the number of affected people by severe climate conditions in Europe dissecting worst events and regional differences. Here, we present a comprehensive analysis of all temperature-related disasters reported in the Emergency Events Database (EM-DAT) for Europe. Ultimately, we propose the development of a working paper for European guidelines for pediatric management of heatwave-related disasters.

## Results

### Overall temperature associated disaster pattern in Europe

Although EM-DAT reports data spanning from 1900 – 2019, it is important to note that data entries before 1988 were retrospectively collected (compare Figure 5). Therefore, we mainly report events following the years of prospective data collection (1988 – 2019), unless stated otherwise.

Since 1988, 367 temperature associated disasters were reported in Europe (see Figure 1). Most events were cold waves (133 = 36.2%), forest fires (80 = 21.8%), and heat waves (69 = 18.8%), followed by severe winter conditions (41 = 11.2%), droughts (36 = 9.8%), land fires (5 = 1.4%), and wildfires (3 = 0.8%, Suppl. Figure 1). Overall, these disasters resulted in 144,650 casualties. Extreme temperature events and its causes in Europe since 1988 were distributed as follows: heat waves (137,533 = 95.1%), cold waves (4,983 = 3.4%), severe winter conditions (1,477 = 1.0%), forest fires (550 = 0.4%), land fires (99 = 0.1%), wildfires (6 = 0,0%), and droughts (2 = 0,0%) (Figure 1, Figure 2). Geographical mapping reveals the distribution of extreme weather-related events that led to high numbers of casualties throughout Europe (Figure 3). For both heat and cold-related events Russia is among the countries with high numbers of casualties. Heat-related casualties were primarily reported in mid-south European countries, whereas only little data is reported in the northern European countries.

**Figure 1.**
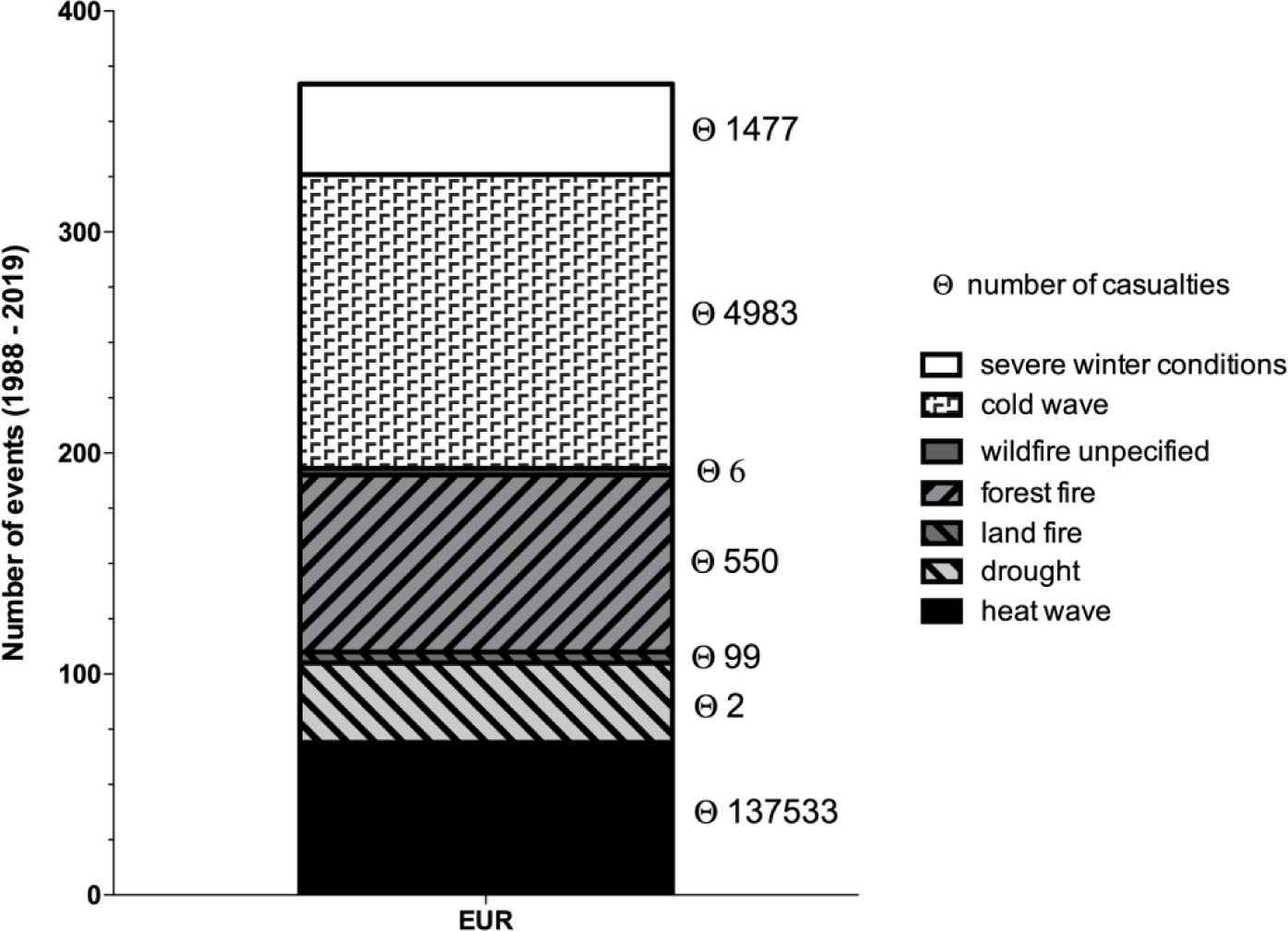
Number of reported disaster subtypes and casualties in Europe between 1988 and 2019. Ɵ = number of casualties

**Figure 2:**
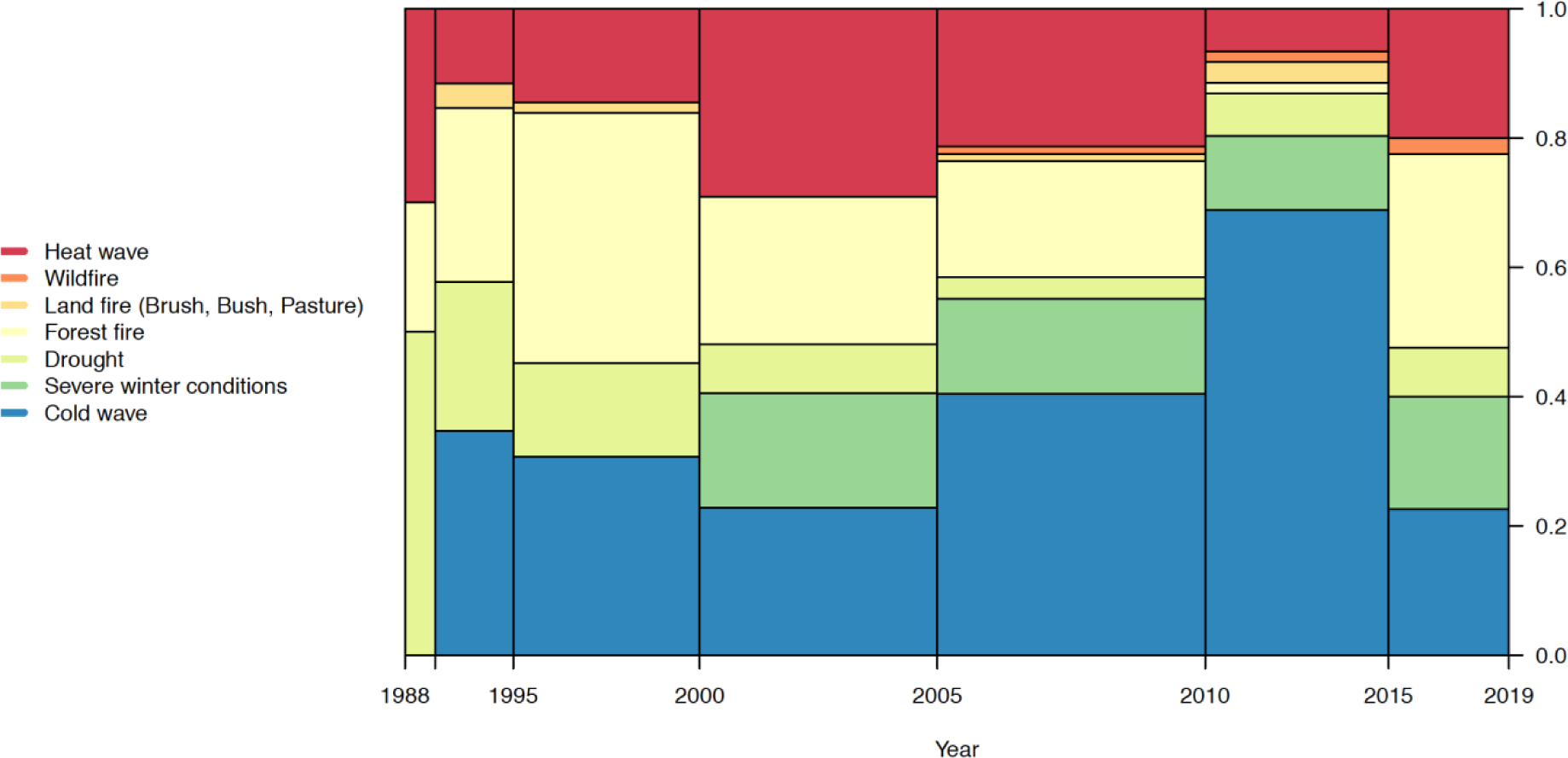
Frequency distributions of temperature associated catastrophic events per 5 year period between 1988 and 2019.

**Figure 3:**
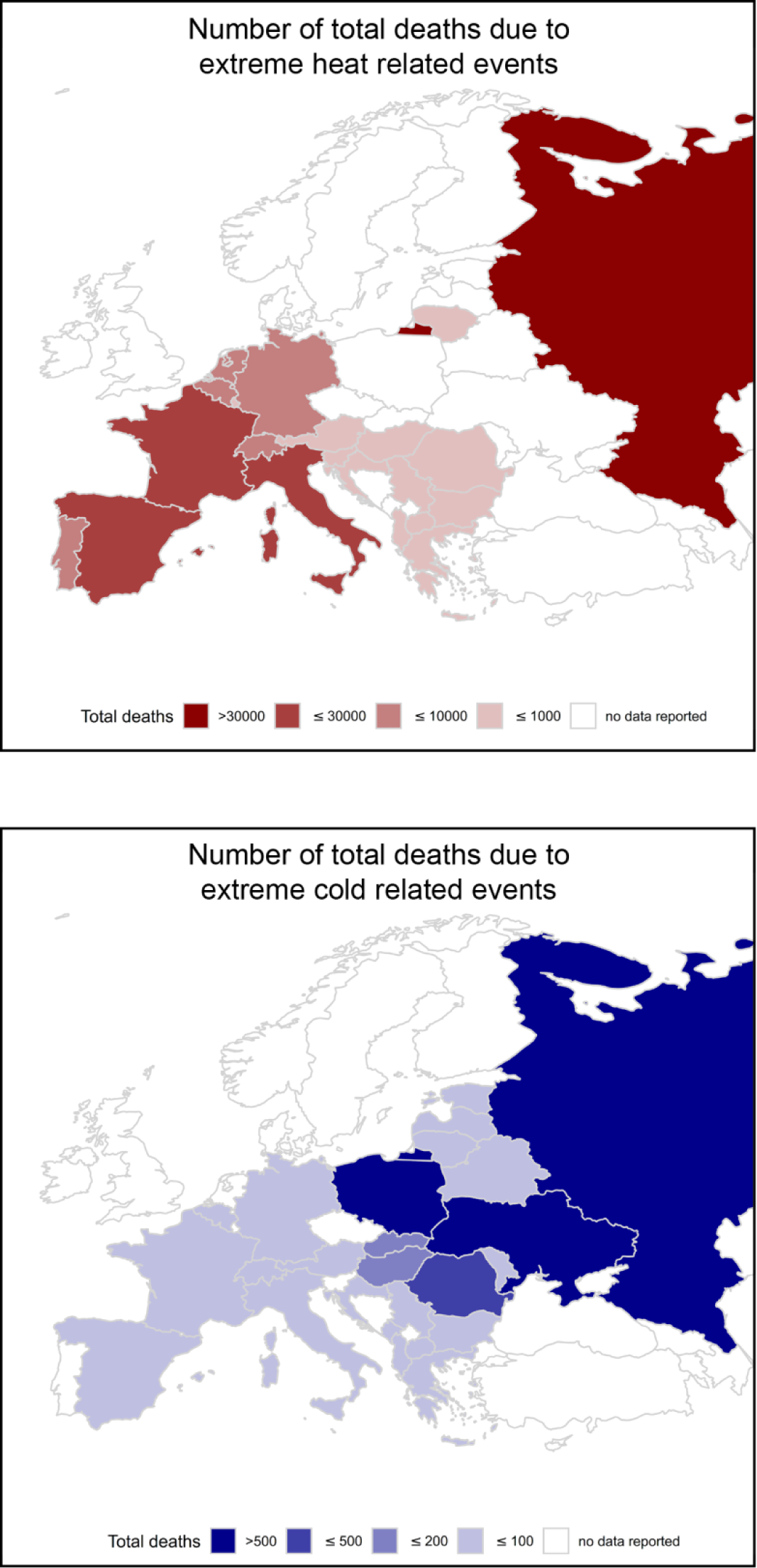
Heatmap of total casualties due to extreme temperature related events throughout Europe during 1988-2019

In the 32 years since 1988, a median of 7 [IQR 4 - 21.25] events occurred per year, most catastrophic events occurred in 2012 (45 = 12.3%), 2000 (29 = 7.9%), 2003 (25 = 6.8%), 2005 (24 = 6.5%), and 2007 (23 = 6.3%). The frequency distribution pattern analysis of temperature associated disasters revealed an increasing number of cold waves until 2015. However, during the period of 2015 to 2019, an overall shift can be observed, characterized by increased numbers of heat waves, and heat-related disasters in EM-DAT (see Figure 2). It is also important to note that during 2005 – 2010 the highest number of events was recorded in EM-DAT (n = 113). In parallel, since 1900, the beginning of the retrospectively collected time period, there have been less records of catastrophic events due to heat waves, yet the difference between casualties due to heat waves versus casualties due to cold waves/severe winter condition was statistically significant (Figure 4 & 5).

**Figure 4:**
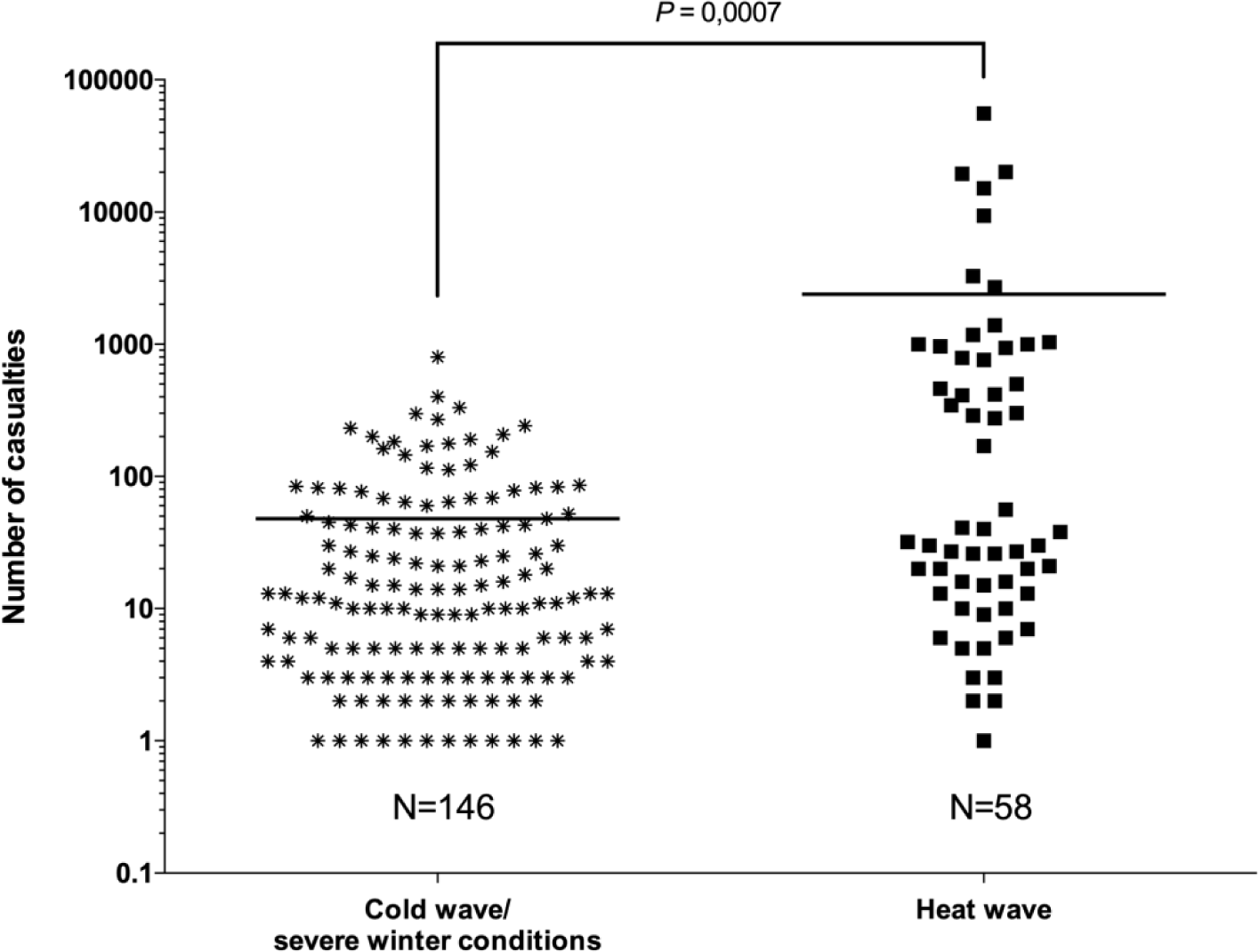
Reported temperature associated casualties in Europe for heat waves and cold weather (cold wave/severe winter conditions) between 1900 and 2019. Horizontal lines indicate means. *p*=.0007 (t-test)

**Figure 5:**
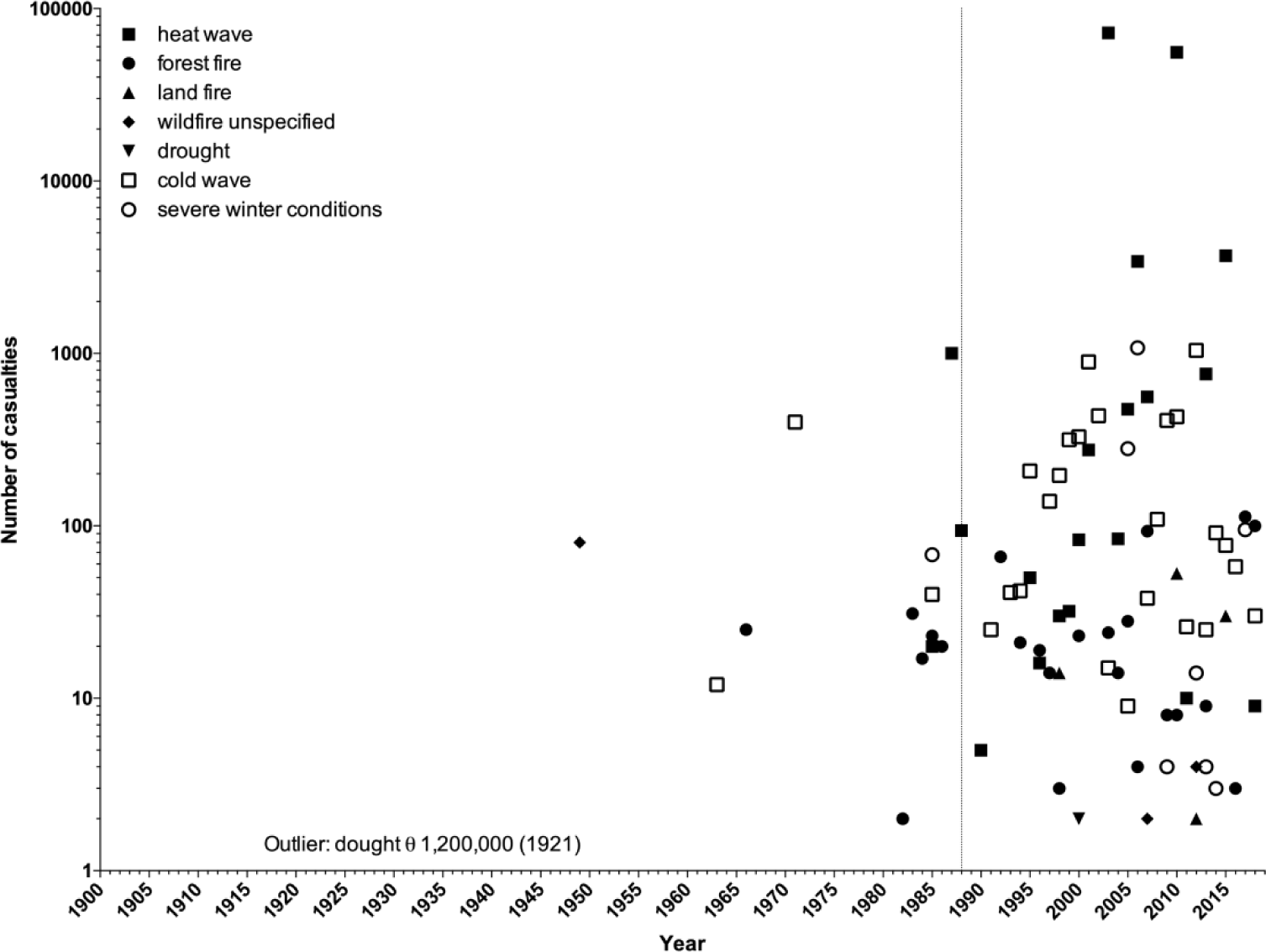
Reported numbers of casualties due to temperature assicated events in Europe by year and subtype between 1900-2019. Ɵ = number of casualties. The vertical line indicates the beginning of prospective data collection in EM-DAT in 1988.The drought in 1921 with 1,200,000 casualties is an outlier and not shown in the graph.

### 5 worst years with pan-European temperature related events for heat waves and cold weather

Since 1900, the five years with most reported casualties due to heat waves in Europe were 1987 (1,000 casualties), 2003 (72,210 casualties), 2006 (3,418 casualties) in 2010 (55,736 casualties), and 2015 (3,685 casualties). In contrast, the five years with the highest number of reported casualties due to cold waves/sever winter conditions were 2001 (894 casualties), 2002 (435 casualties), 2006 (1,077 casualties), 2010 (429 casualties), and 2012 (1,042 casualties) (Figure 5).

### Worst heat-related events in Europe

Catastrophic heat waves with more than 100 casualties are listed in Table 1. The overall top ten of twenty-five recorded heat waves with more than 100 casualties in Europe happened during the last 15 years with a total number of 129,333 casualties, whereas the number of casualties due to cold related events was 3,174, thereby significantly lower, and no accumulation of event frequency was observed.

**Table 1:** Reported catastrophic heat waves with more than 100 casualties in Europe by country, year, and temperature between 1900 and 2019.

| Country | Year | Temperature<br>magnitude value [°C] | Casualties |
| --- | --- | --- | --- |
| Russia | 2010 | 40 | 55736 |
| Italy | 2003 | NA | 20089 |
| France | 2003 | 43 | 19490 |
| Spain | 2003 | 40 | 15090 |
| Germany | 2003 | NA | 9355 |
| France | 2015 | 40 | 3275 |
| Portugal | 2003 | NA | 2696 |
| France | 2006 | 37 | 1388 |
| Belgium | 2003 | NA | 1175 |
| Switzerland | 2003 | NA | 1039 |
| Greece | 1987 | 41 | 1000 |
| Netherlands (the) | 2006 | 34 | 1000 |
| Netherlands | 2003 | NA | 965 |
| Belgium | 2006 | NA | 940 |
| Croatia | 2003 | NA | 788 |
| United Kingdom of<br>Great Britain (the) | 2013 | 40 | 760 |
| Hungary | 2007 | 41.9 | 500 |
| Portugal | 2005 | NA | 462 |
| Czechia | 2003 | NA | 418 |
| Belgium | 2015 | NA | 410 |
| Austria | 2003 | NA | 345 |
| United Kingdom of<br>Great Britain | 2003 | NA | 301 |
| Slovenia | 2003 | NA | 289 |
| Russia | 2001 | 30 | 276 |
| Luxembourg | 2003 | NA | 170 |

Missing values indicate that no data was reported to EM-DAT. Data before 1988 were collected retrospectively.
NA – not applicable

### Worst cold-related events in Europe

European catastrophic cold waves and severe winters with more than 100 casualties are shown in Table 2, cold waves and severe winter with more than 1000 individuals affected are listed in Table 3.

**Table 2:** Reported catastrophic cold waves and severe winter conditions with more than 100 casualties in Europe by country, year, disaster subtype and temperature between 1900 and 2019

| Country | Year | Disaster subtype | Temperature magnitude value [°C] | Casualties |
| --- | --- | --- | --- | --- |
| Ukraine | 2006 | Severe winter conditions | -30 | 801 |
| Spain | 1971 | Cold wave | NA | 400 |
| Russia | 2001 | Cold wave | -26 | 332 |
| Poland | 2009 | Cold wave | -35 | 298 |
| Poland | 2001 | Cold wave | -25 | 270 |
| Russia | 2002 | Cold wave | -50 | 242 |
| Russia | 2000 | Cold wave | -7 | 232 |
| Russia | 1995 | Cold wave | NA | 208 |
| Poland | 2010 | Cold wave | -33 | 200 |
| Poland | 2005 | Severe winter conditions | -32 | 191 |
| Poland | 2002 | Cold wave | -25 | 183 |
| Poland | 2012 | Cold wave | NA | 177 |
| Russia | 2012 | Cold wave | NA | 170 |
| Russia | 1999 | Cold wave | NA | 162 |
| Poland | 1999 | Cold wave | -27 | 154 |
| Russia | 2001 | Cold wave | -57 | 145 |
| Slovakia | 2010 | Cold wave | NA | 122 |
| Russia | 2006 | Severe winter conditions | -43 | 116 |
| Ukraine | 2012 | Cold wave | -30 | 112 |

Missing values indicated that no data was reported to EM-DAT. Data before 1988 were collected retrospectively.
NA – not applicable

**Table 3:** Reported catastrophic cold waves and severe winter conditions with more than 1000 people affected in Europe by country, year, disaster subtype and temperature between 1900 and 2019

| **Country** | **Year** | **Disaster subtype** | **Temperature magnitude value [°C]** | **Total population affected** |
| --- | --- | --- | --- | --- |
| Russia | 1999 | Cold wave | -50 | 725000 |
| Albania | 2012 | Severe winter conditions | NA | 230000 |
| Ukraine | 2012 | Cold wave | -30 | 87500 |
| Serbia | 2012 | Cold wave | -30 | 70000 |
| Ukraine | 2006 | Severe winter conditions | -30 | 59600 |
| Belarus | 2017 | Severe winter conditions | -31 | 50539 |
| Slovenia | 2014 | Severe winter conditions | NA | 50000 |
| Belarus | 2014 | Severe winter conditions | -35 | 31500 |
| Russia | 2002 | Cold wave | -50 | 25062 |
| Serbia | 2012 | Severe winter conditions | NA | 18234 |
| Belarus | 2013 | Severe winter conditions | -25 | 11325 |
| Bosnia and Herzegovina | 2012 | Cold wave | -32.5 | 10347 |
| France | 1997 | Cold wave | NA | 10000 |
| Bosnia and Herzegovina | 2009 | Severe winter conditions | -25 | 10000 |

Missing values indicated that no data was reported to EM-DAT. Data before 1988 were collected retrospectively.
NA – not applicable

### Casualties by magnitude

Figure 6 shows casualties by temperature magnitude for catastrophic cold-waves and severe winters versus heat waves. Casualties for extreme low temperature were observed at temperatures between – 7 °C and – 57 °C, whereas casualties due to extreme heat were at temperatures between 30 °C and 47 °C. Casualties at a given temperature varied intensely and we could not detect any temperature magnitude - number of casualties dose – effect - relationship. The highest number of extreme-cold associated casualties occurred in the Ukraine (801 casualties), whereas the highest casualties for heat-associated casualties were observed in Russia (55,736 casualties), Italy (20,089 casualties), France (19,490 casualties), and Spain (15,090 casualties) (see Table 1 & 2).

**Figure 6:**
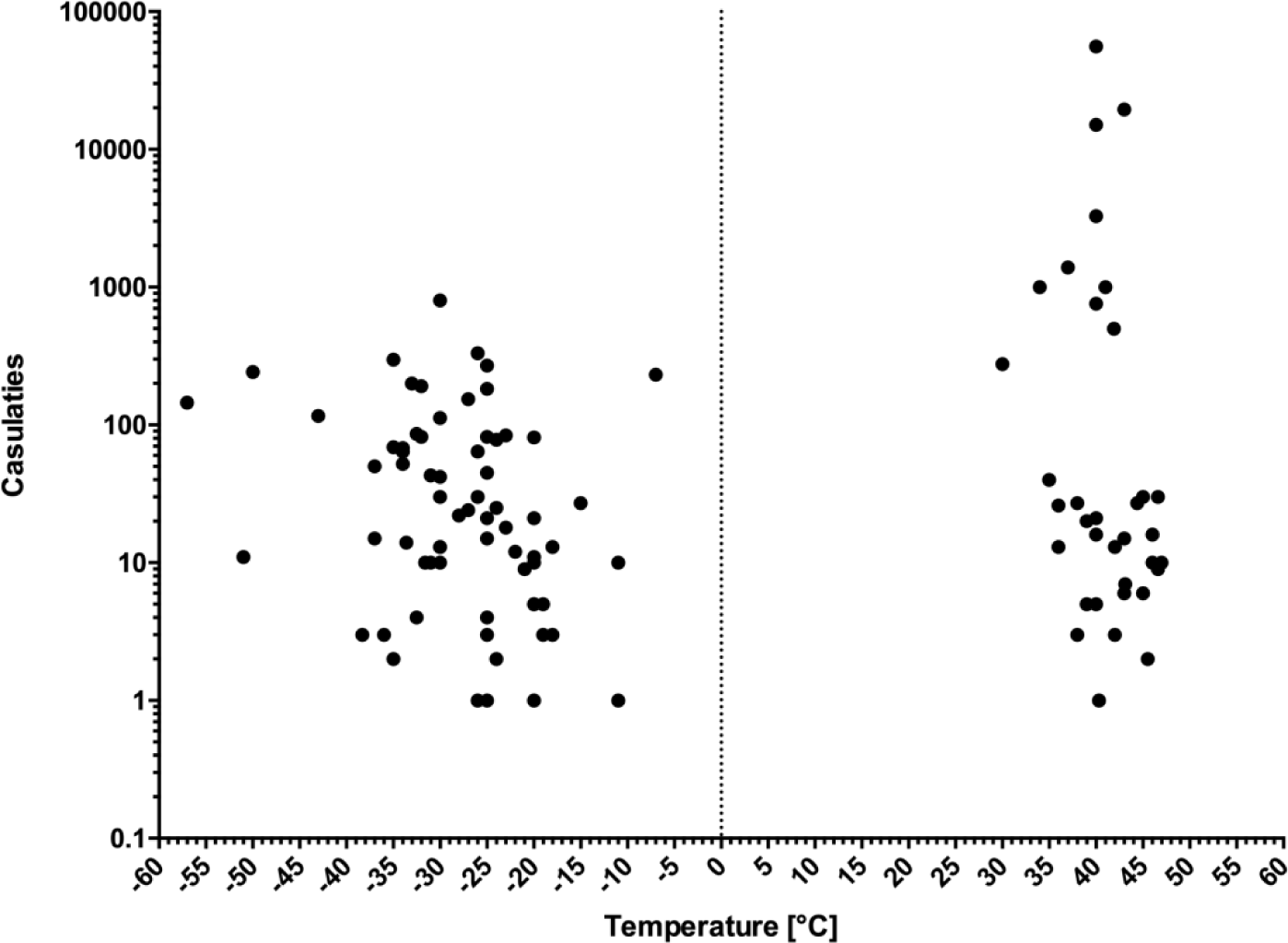
Casualties by temperature magnitude for catastrophic temperature related events, i.e. heat waves and cold waves / sever winter conditions in Europe between 1900 and 2019.

### Change of temperature in Europe over time

We analyzed temperature data provided by the Worldbank Climate Change Knowledge Portal (Figure 7 & Suppl. Figure 2). This data demonstrates rising average surface temperatures in all 49 European countries. A stronger trend can be observed from 1975 onwards.

**Figure 7:**
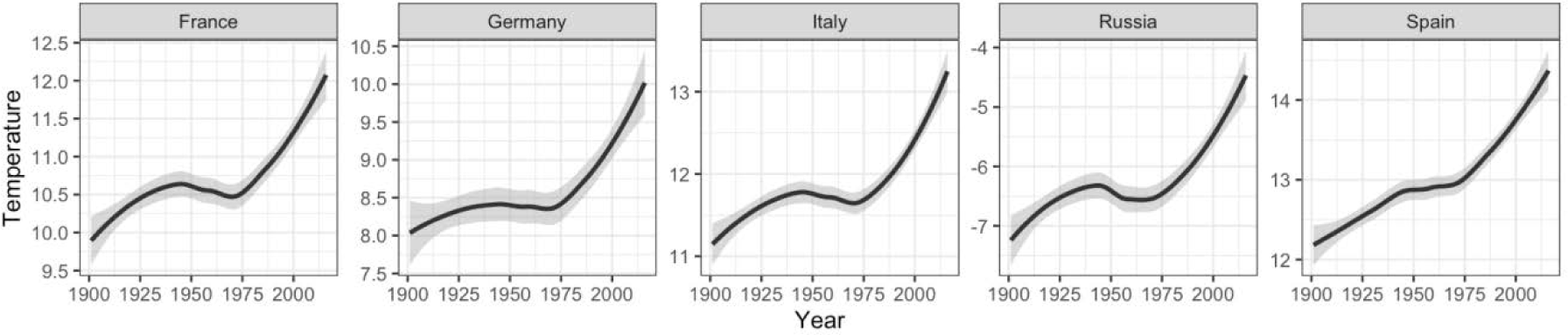
Average surface temperature development during the years 1900 – 2016. Depicted is a subset of countries (France, Germany, Italy, Russia, and Spain) that was affected most severely by heat waves. For a full display of all European countries see Suppl. Figure 2.

## Discussion

Heatwaves, although less frequently reported than cold waves or severe winter conditions, were associated with significantly more casualties in Europe than cold temperature extremes. This confirms and extends a previous analysis of data from Germany and France to a pan-European level.^5^ The present data suggest that heat waves are a serious European public health threat. For the five deadliest years for heat waves in Europe, i.e., 1987, 2003, 2006, 2010, and 2015, a total of 136,049 casualties were reported in EM-DAT (Figure 1 & 4, Table 1). The five deadliest European heat waves on a country level occurred in Russia (2010) as well as Italy, France, Spain, and Germany (all in 2003) and led to 119,760 reported casualties in these five countries (Table 1). Thus, the 2003 heat wave represents one of the most serious and significant natural disasters due to climate phenomena in the last 100 years of European history.

Heatwaves are commonly referred to as heat or hot weather that lasts for several days. According to the World Meteorological Organization, the criteria of a heat wave are met, when for a specific region during five or more consecutive days the maximum daily temperatures are at least 5°C above the average temperature for that time of year. Current heat waves are attributed to changes of the jet stream over Europe. Blocks, defined as large-scale high pressure systems, prevent the formation of clouds, thereby allowing more radiation to reach the surface of the planet and increase temperature.^7-9^ Heat waves can usually be anticipated within a time frame of days to months,^10^ although heat event definitions and threshold for triggering warnings vary within jurisdictions in Europe and the rest of the world.^11^

The European climate ranges from subtropical in the southern countries to temperate in the middle and northern parts. Over the last 100 years, the average surface temperature has been continuously rising. During winter months, cold days occur less frequently, while heat waves present with a higher frequency, longer duration, and overall increased intensity.^12^

We observed an overall rise in average surface temperature for all European countries within the last 100 years independent of their geographical distribution. This trend seems to be markedly enhanced since 1975 (Figure 7 & Suppl. Figure 2). In parallel to this observation, numbers of extreme temperature-related casualties have been rising within this observational period. We expect a strong impact of rising temperatures on the overall health of the pediatric population. Due to inherent physiological properties, children represent a particularly vulnerable population in regard to temperature related health threats: higher metabolic rate,^13^ higher surface area,^3^ smaller absolute blood volume,^14^ lower rate of sweat production.^13,15^ Excessive heat causes cellular injury through mechanisms of inflammation, and protein denaturation,^16,17^ but also through direct injury of vascular architecture and impaired microcirculation.^18^ Symptoms of hyperthermia-related illness appear when the body’s ability to regulate temperature are overwhelmed and include excessive sweating, thirst, tachycardia, nausea, vomiting, neurologic symptoms, including headaches and behavioral changes, seizures, delirium, or coma.

Excessive heat has also been linked to considerable negative effects on mental health. It has been shown to increase psychiatric hospitalization rates and prevalences of psychiatric disorders, predominantly depressive disorders, anxiety, insomnia and substance use disorders.^19-24^ Mediated by elevated stress hormone release, excessive heat can also lead to more aggressive behaviors and higher crime rates.^25-27^ As in other types of disasters, traumatic events caused by temperature related disasters may lead to adjustment disorder or posttraumatic stress disorder in both adults and children.^28,29^ Yet, the development of a psychiatric disorder after experiencing a disaster does not occur by default in all individuals.^30^ This can be attributed to the construct of psychological resilience, which describes the ability to overcome adverse situations without negative long-term effects. Therefore, strengthening resilience by effective measures before, during and after disasters (as proposed in Table 4) as well as identifying vulnerable populations at risk represent viable strategies in preventing disaster related psychiatric disorders. Furthermore, persons with psychiatric disorders represent another particularly vulnerable population in regard to temperature related health threats.^31,32^ Consequently, mentally ill children are burdened with a combined risk of suffering from negative outcomes caused by temperature related disasters.

**Table 4:** Pediatric crisis measures and mitigation strategies for heat waves

| **Purpose** | **Measure** |
| --- | --- |
| Ensure sufficient fluid intake (drinking) | • Increase fluid intake without waiting to feel thirsty<br>• Raise awareness to caregivers to be alert to hydration levels in children, in particular bedridden or cognitively impaired individuals |
| Avoid increase of body temperature (cooling) | • Establish cooling centers<br>• Ensure intuitive and transparent awareness of localization and opening times and stock with first aid materials, drinking water, games or activities for children<br>• Establish spray parks and provide cooling services for children, ranging from toddlers to teens. |
| Educate caregivers (schools and childcare providers) | • Ensure that adequate cooling measures are in place<br>• Avoid strenuous activities during the hottest part of the day<br>• Educate children on actions to reduce heat risks, which they can then share at home<br>• Ensure that schoolchildren have adequate access to water and cool areas to rest |
| Protect vulnerable populations | • Organize home outreach visits to vulnerable people<br>• Evacuate vulnerable people from their homes to cooling centers |
| Ensure medication safety for individuals with chronic health conditions | • Consider drug interference with sweating, thermoregulation, thirst, hydration, electrolyte balance, renal function, blood pressure, level of alertness, and potentially increased toxicity due to dehydration<br>• Check if medications need to be stored in a refrigerator |

[In reference to: Singh, R., Arrighi, J., Jjemba, E., Strachan, K., Spires, M., Kadihasanoglu, A., Heatwave Guide for Cities. 2019. Red Cross Red Crescent Climate Centre p-18, accessed 26 July 2019,<sup>63</sup> and Beat the Heat - Information For Health Professionals,<sup>66</sup> accessed 26 July 2019]

Disaster risk perception in the population may not reflect the situation appropriately and either over- or underestimate true possibilities for disasters. Although between 1900 and 2009, EM-DAT lists 60,200 casualties associated for heat waves in Europe (Figure 4 & 5), the 2009 Special Eurobarameter civil protection survey conducted in 27 European Union member states covering 26,470 interviews out of a population size of 406,557,138 individuals over 15 years, listed flooding (45%), violent storms (40%), industrial accidents (29%), forest fires (27%), earthquakes (22%), marine pollution (20%), nuclear accidents (16%), landslides (7%), volcano eruptions (2%), tsunamis (1%), others (1%), no answer (2%), and none (8%). Heat waves were not being mentioned (https://ec.europa.eu/commfrontoffice/publicopinion/archives/ebs/ebs_328_en.pdfpage6, accessed 06 Sept 2019). Therefore, given the data in this report, it is imperative to educate the European public about the danger of heat waves in order to appropriately 1) prevent 2) respond to, and 3) recover from heat related disaster. As such, possible pediatric crisis measures and mitigation strategies for heat waves are listed in Table 4.

A concept of improved adaptation to cold temperature might be found in view of evolutionary developmental aspects: Human resilience towards heat seems to have evolved already in early members of the genus *Homo*, when the body’s ability to adapt its metabolism according to surrounding temperature emerged to represent a major advantage.^33-36^ In contrast, tolerance towards cold is frequently dependent on behavioral support, such as clothing, equipment, and shelter.^37^ From an evolutionary point of view, our species originated in the hot climates of Africa between 300,000-200,000 years ago,^38,39^ with the typical linear physique (tall and lean) of early *Homo sapiens* clearly indicative of heat-related adaptations.^36,40^ Recent archaeological studies of our deep history between 40,000-10,000 years show that as *Homo sapiens* was expanding around the globe, and well before the origins of agriculture and a sedentary lifestyle, these populations were able to inhabit both high-altitude and cold places such as the Peruvian Andes, the glacial Bale mountains of Ethiopia and Arctic Siberia^41-43^ but also hot rainforest and desert environments.^44,45^ Some concomitant physiological adaptations to extreme temperatures throughout the last 100,000 years are also indicated by DNA, aDNA and anatomical research.^46-49^ Migrating into Europe by ∼40,000 years ago, *Homo sapiens* faced the Neanderthals, which show clear anatomical adaptations to cold stress and more temperate climates in their postcranial and cranial features.^40,50,51^ Yet, modern humans outcompeted Neanderthals in their homeland, suggesting that originally heat-adapted members of our species were able to quickly adapt to cold stress encountered in these novel environments. The reasons are likely found in advanced cultural innovations such as the ability to make fire, as well as specialized cold weather clothing and hunting weapons.^52-55^ Archaeological remains over the past 40,000 years in Europe show that these “cultural buffers”^48,56^ increase exponentially through time – with sheltered houses, herbal medicines and organized water supply in the Holocene as some of many examples – aiding in both cold and heat extremes. A hypothesis deriving from this deep-time evolutionary perspective is that increasing reliance on behavioral solutions and cultural innovations to extreme temperatures led to a relaxation of selection pressures on anatomical, genetic and physiological adaptations (see also ^48^), particularly in European regions where these events happened rarely. In pre-industrial times, existing behavioral adaptations to extreme cold were more efficient than those to extreme heat: adding layers of clothes and generating more heat is always possible, whereas further cooling down the naked human body presents a more difficult challenge.

Today, the central European population has widely access to mentioned cold temperature behavioral support structures, while the number of households with e.g. air conditioning is reported to be below 5%. (https://webstore.iea.org/the-future-of-cooling; Report available). In line with this observation, the presented data may indicate that the population resilience against cold is much higher than it is against heat.

Catastrophic heat wave events with high casualties tended to occur rather in Western/Southern Europe (exception: Russia) whereas cold waves/severe winter conditions with casualties tended to focus on Central and Eastern Europe (Tables 1 and 2, Figure 3). The highest number of cold-related casualties was observed in Ukraine, a country with mostly temperate climate. However, due to the Siberian Anticyclone effect, temperatures can drop to -30 °C.^57^ Interestingly, heat-associated casualties occurred mainly within Central/Western Europe (France, Spain, The Netherlands) (Table 1, Figure 3). The 2003 heat wave is explained by researchers through a stationary ridge over western Europe.^58^ The abundance of heat-wave related casualties might be attributed to the missing awareness of the danger of such of the people in central and Western Europe and illustrates the need for national and international campaigns to raise awareness.

Year after year, extreme temperature records are broken as climate change causes an incline in heat-wave frequency and length in Europe: out of all records, the 18 highest average temperatures recorded during summer months occurred over the last 19 years. Hot summers are associated with an increased risk of fire which may require evacuation measures and can lead to casualties (Figure 1, Figure 6).

Besides its direct effect on health and viability, extreme temperature also causes a major threat to the agricultural sector. Drought is defined as a climate-associated period of low precipitation and high temperatures, which can lead to substantial damage of agricultural production across the globe.^59^ For many developing countries, droughts represent the major hazard effector of damage and loss in the agricultural sector, especially in regard to livestock. In Australia, droughts have been linked to higher suicide rates in farmers.^60,61^ However, recent droughts in Europe associated with heat waves and a lack of precipitation also cause socio-economical damage to a majority of European countries.^62^ After a drought in Russia in 1921 with associated 1,200,000 casualties due to crop failure and famine, resilience to droughts seem to have improved in Europe, because 44 more droughts were reported thereafter (between 1976 and 2018) and only one drought event was reported with reported 2 casualties in the Republic of Moldova in 2000 (data not shown). It is likely that the awareness of drought periods is much higher due to its immediate effect on agricultural goods and indirect consequences for viability. Measures taken to prevent crop failures or loss of livestock due to droughts are manifold, including improvement of agricultural methods such as reduction of water with low-flow appliances, selection of heat-resistant crops, and surveillance of livestock. National and international task forces are in place to continuously monitor the changes observed in regard agricultural yield. Comparable measures and awareness-raising campaigns on a pan-European level would likewise help to tackle the increasingly pressing threat of heat waves.

Unfortunately, population data in EM-DAT are not stratified by age, which would allow a more targeted approach towards disaster epidemiology.^5^ Pediatric populations particular vulnerable to heat include children under five years, adolescents working outside (e.g. in a professional apprenticeship), individuals with chronic health conditions, obese individuals, migrants and refugees, and children of illiterate or non-native language speaking families.^63^

### Limitations and directions for future research

As previously outlined, the analysis of EM-DAT has some important limitations which have to be taken into account for the appropriate interpretation of the present findings.^5^ First, the analysis relies on the accuracy of the reported data. Second, retrospective data, i.e. events before establishing the database in 1988 may be incompletely registered and we cannot rule out reporting bias. EM_DAT does not capture data related to warfare which may further contribute to an underestimation of temperature associated casualties in the past. This probable ascertainment bias is illustrated in Figure 5 where in particular for the two major events of warfare and destruction in Europe during 1914-1918 and 1939-1945, no temperature associated casualties are reported in EM-DAT which is unlikely reflecting the true situation. Nevertheless, although likely incomplete, the retrospective information in the database is of importance such as the data entry drought in the Soviet Union in 1921 with a major impact on the population. Missing data were not imputed. The differences in data density between the retrospective and prospective data collection periods do not indicate an increase of catastrophic events between the two periods, but are rather due to ascertainment bias and data availability. Fourth, epidemiological surveillance in each single country included in the present analysis may vary which may introduce data heterogeneity. Fifth, temperature magnitude for a particular catastrophe is coded as a single number in the database, may not be necessarily representative for the overall characteristics of the extreme temperature event, as it does not consider a) the length of the event or b) particular local geography. Sixth, as the largest country by size, Russia is represented as a single country in EM-DAT. The available data did not allow for a more precise geographical analysis of temperature related disasters, e.g. in which region of Russia (Europe vs. Asian) a disaster took place. Seventh, data in EM-DAT are not age stratified which would allow a more detailed analysis of vulnerable populations including children and facilitate better data-driven pediatric decision making in catastrophic situations in the future. Nevertheless, EM-DAT is an authoritative state of the art data source,^5^ and we consider the present findings generalizable within the context of the previously described limitations.

### Conclusion

In this report, we show that according to EM-DAT data, extreme temperatures are an increasingly significant public health threat to the European population as the average European surface temperatures are increasing. Although cold waves are more frequently reported in EM-DAT, heat waves are the major cause for temperature-related casualties. Therefore, we conclude that evolutional and cultural resilience against cold and drought is significantly higher than it is against heat and explain evolutionary concepts behind this hypothesis. It is projected that the frequency, duration and intensity of heat waves is to further increase due to current climatic changes and will therefore become an even more prevalent problem for future generations. Therefore, we see a need to formulate an emergency plan to inform the public and authorities about measurements to be taken in such extreme heat conditions to conquer the prevailing lack of information available to the public.

## Methods

### Endpoints

Primary endpoints were: Reported events and casualties associated with extreme temperatures (heat waves, cold waves, severe winter conditions) as well as droughts and wildfires. Secondary endpoints were number of individuals affected by droughts, cold waves, and severe winter conditions. Tertiary, i.e. explorative endpoints were geographical differences, changes over time, juxtaposition to mean temperature. Main focus of the present analysis was the prospectively collected data since database launch in 1988, the retrospectively reported data in EM-DAT covering the time span between 1900 to 1987 were considered exploratory.

### Data source

Similar to a previous epidemiological study of disasters in Germany and France,^5^ the present data analysis based on EM-DAT (Emergency Events database), launched in 1988, maintained by the Centre for Research on the Epidemiology of Disasters (CRED) at the School of Public Health of the Université catholique de Louvain located in Brussels, Belgium [https://www.emdat.be/frequently-asked-questions, accessed 12 July 2019]. EM-DAT captures data on more than 21,000 disasters worldwide between 1900 and today, data sources are UN agencies, governments, the International Federation of Red Cross and Red Crescent Societies, other non-governmental organizations, insurance companies, research institutes and press agencies [https://www.emdat.be/frequently-asked-questions, accessed 12 July 2019]. Inclusion criteria for a disaster the Emergency Events database are one or more of the following criteria fulfilled: (1) 10 or more people dead, (2) 100 or more people affected, (3) the declaration of a state of emergency, or (4) a call for international assistance [https://www.emdat.be/frequently-asked-questions, accessed 12 July 2019]. Suppl. Figure 1 was created with the application SnapPlanet (https://snapplanet.io/) using European Space Agency Sentinel-2 satellite imagery.

### Data query

EM-DAT (https://www.emdat.be/) was accessed over the internet on July 10^th^, 2019. The following advanced search parameters were applied: period: 1900 to 2019; location: continent Europe (as documented by EM-D https://www.emdat.be/guidelines, i.e. according the UN regional division https://unstats.un.org/unsd/methodology/m49/); disasters classification: climatological: drought, wildfire (unspecified, forest fire, land fire); meteorological: extreme temperature (cold wave, heat wave, severe winter conditions). The following variables were extracted and analyzed: catastrophic evet start date, country, disaster type, disaster subtype, total casualties, total affected, temperature magnitudes. The queried dataset was transmitted to MR as validated data though email by EM-DAT on July 12^th^, 2019. Last database update was April 19^th^, 2019. Data were manually checked for plausibility. For geographical mapping (Figure 2) events were attributed to the precise regional localization according to current geographical jurisdictions (e.g. “German Federal Republic – DFR” was recoded to “Germany – DEU”). Missing data were not imputed.

The World Bank Climate Change Knowledge Portal was accessed on July 3^rd^, 2019. Monthly surface temperature data for all European countries from 1901-2016 was downloaded and used to calculate a yearly average temperature per country. Average yearly temperatures were plotted using an additive model of measurements using local polynomial regression fitting in RStudio (Version 1.2.1335).

### Statistical analysis

Strengthening the reporting of observational studies in epidemiology (STROBE) criteria, a quality assurance reporting guideline and checklist for observations studies, were respected.^64^ As the EM-DAT started collecting data prospectively in 1988, the time period for the primary and secondary endpoints was set between 1 January 1988 until last database update before data download (i.e., 19 April 2019) in order to accommodate reporting and ascertainment bias. Standard methods of descriptive statistics were applied. Variables were summarized using counts and percentages of the total study population.

Differences between casualties associated with heat waves vs. cold waves/severe winter conditions were calculated with standard t-test procedures. A two-sided P-value of equal or less than .05 was considered statistically significant.

In order to detect meaningful biological and epidemiological differences between vulnerabilities towards heat and cold, 1) number of disaster events, 2) number of casualties, and 3) numbers of persons affected were compared between a) heat waves and b) cold waves and severe winter conditions, similar to the analysis conducted for Germany and France before.^5^ The analysis of EM-DAT data was conducted with GraphPad PRISM 5.04 (La Jolla, CA, USA), and R (http://www.r-project.org). The European map was plotted using the R extension “ggmap”.^65^

## Data Availability

All data generated or analyzed during this study are included in this article (and its Supplementary Information files).

## Acknowledgments

H.B. received support from the Physician-Scientist Program at Ruprecht-Karls-University Heidelberg Faculty of Medicine.

## Author contributions

H.B. and M.R. developed the concept of the survey and formulated hypotheses. H.B., M.R., E.R, S.G. organized and analyzed the data. S.G. performed all statistical analyses. H.B., M.W., K.M., S.G., and M.R. helped interpreting the results. All authors provided critical feedback and helped shape the research, analysis and manuscript.

## Data availability

All data generated or analyzed during this study are included in this published article (and its Supplementary Information files).

## Figures

**Suppl. Figure 1:**
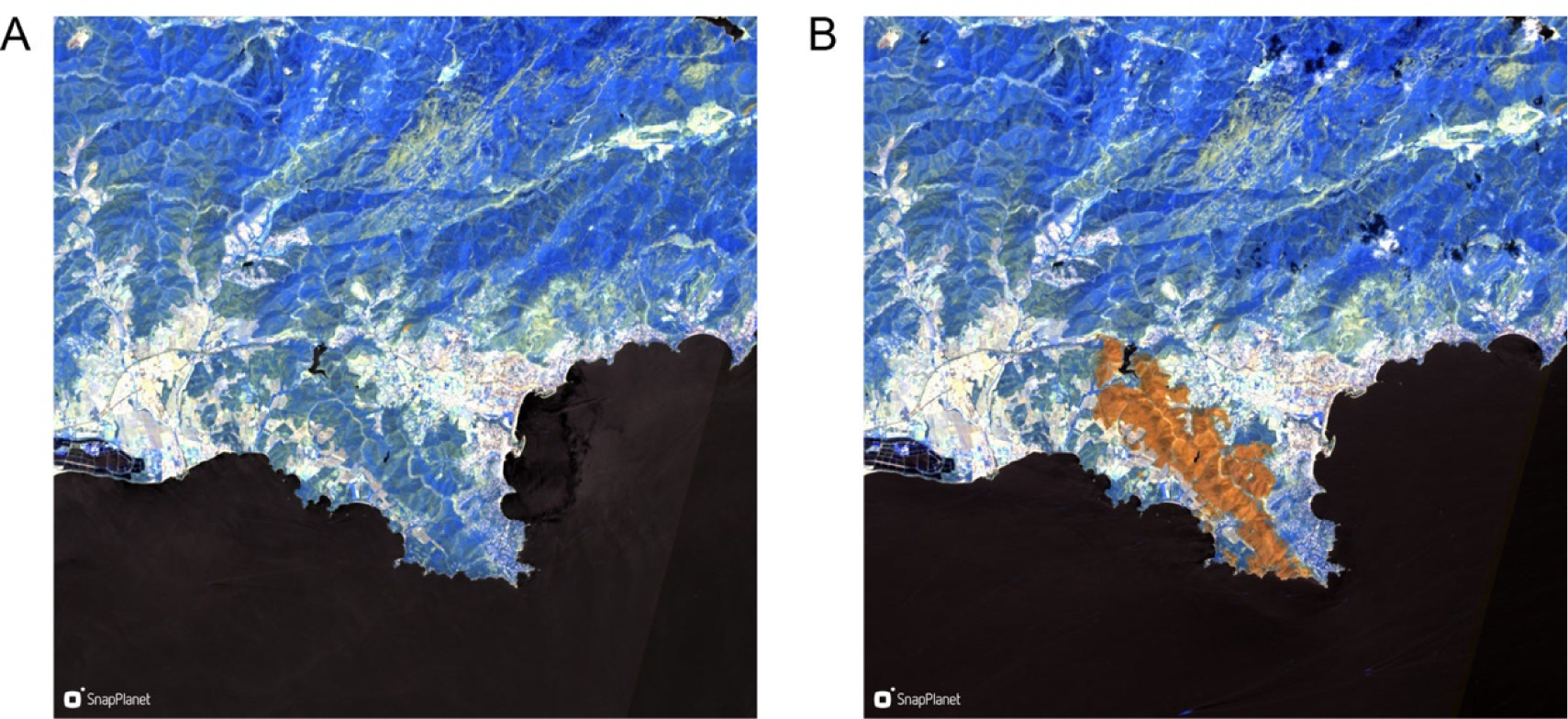
Wildfire near Bormes-les-Mimosas (Var-cote d’Azur, France), 24-25 July 2017, SnapPlanet Sentinel-2 infrared satellite image before (A, 14 July 2017) and after (B, 29 July 2019). The burnt area in the post-fire image appears in orange. This disaster inluded additional areas in Haute-Corse, Vaucluse, Var, and Alpes-Maritimes affecting 12,012 people overall.

**Suppl. Figure 2:**
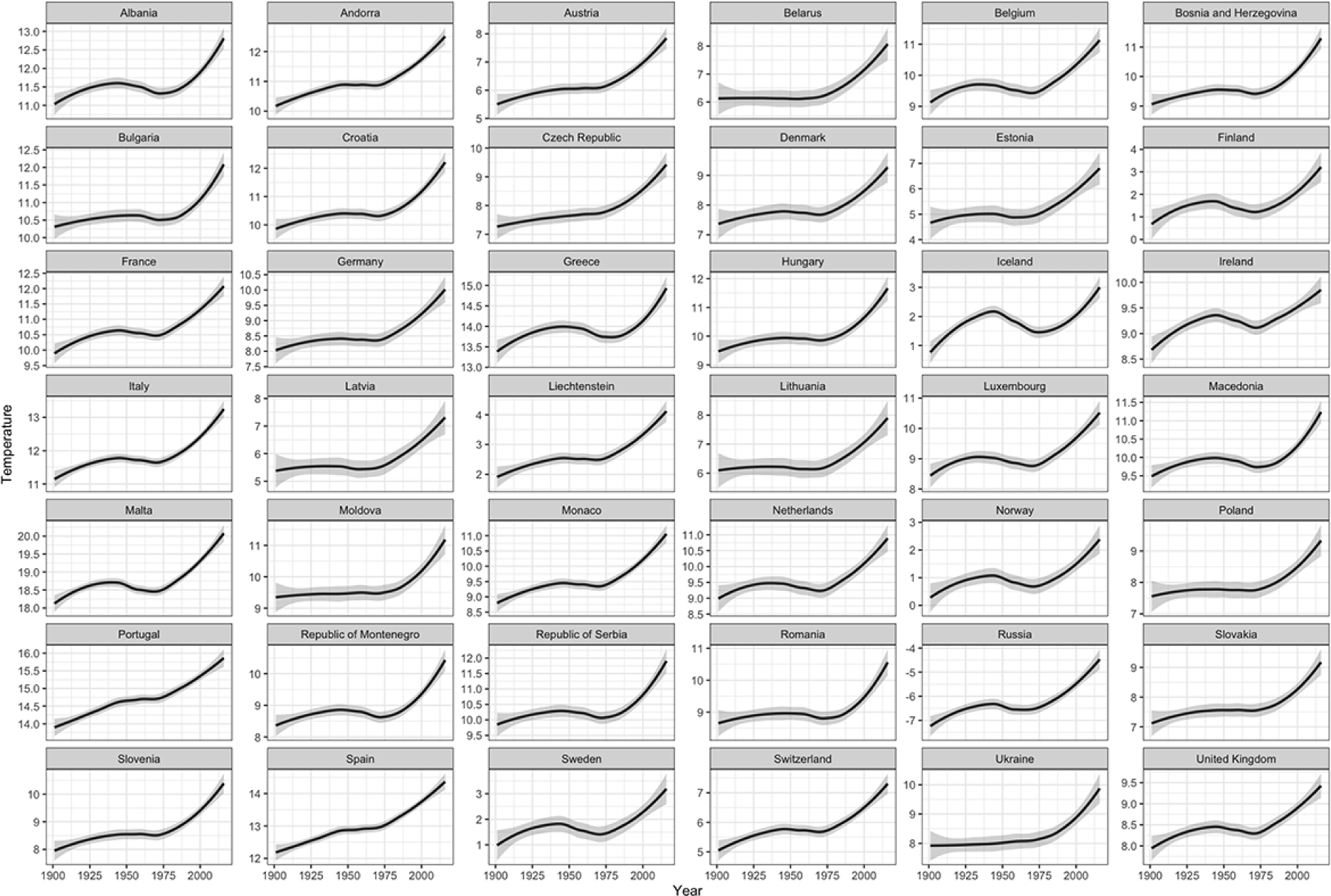
Average surface temperature of 47 European countries over a time period of 116 years. A generalized additive model was built using the geom_smooth function by a quadratically penalized likelihood type approach to generate a smooth line with confidence intervals.

